# Nurses’ perceptions of videoconferencing telenursing: comparing frontal learning vs. online learning before and after the COVID-19 pandemic

**DOI:** 10.1101/2023.05.22.23290291

**Authors:** Svetlana Kats, Liora Shmueli

## Abstract

**Introduction:** During the COVID-19 pandemic, a digital transformation led to an expansion in telenursing practices and a shift in training to online learning. The aim of this study was to explore the impact of behavioral-related factors, based on both TAM and TPB variables, on the intention to use telenursing through videoconferencing and to compare the effect of frontal (before COVID-19) vs. online (during and after COVID-19) training in post-basic nursing courses on nursing attitudes to telenursing.

**Methods:** A cross-sectional online survey was conducted in December 2022 among nurses working mainly at hospitals in Israel who underwent post-basic education training between January 2017 and December 2022. A multivariate ordinary least squares (OLS) regression analysis was used to investigate determinants of intention to use telenursing through videoconferencing

**Results:** Nurses have a positive attitude towards telenursing technology via videoconferencing for remote patient care, regardless of whether they learned about it through face-to-face or online training. The ease of use and the perception of the technology’s importance by colleagues and supervisors were found to have the most significant impact on the attitude of both research groups towards the use of telelearning.

**Discussion:** Successful implementation of new technology in healthcare requires organizational and collegial support. Therefore, managers should encourage the use of telenursing by providing appropriate training for nurses and the necessary resources and support.

## 1. Introduction

The COVID-19 pandemic accelerated digital transformations that required adjustments in occupational settings and training among medical teams, especially among nurses, who constitute the largest healthcare workforce. This period was marked by the mass adoption by healthcare providers worldwide of digital technologies and technology-based virtual platforms (i.e., telehealth and telenursing) aimed at prevention and monitoring. Consequently, the COVID-19 pandemic has set a new standard of care for nurses and has changed their workforce roles [1]. An open question in the field is whether the increased use of remote technologies due to the COVID-19 pandemic has changed nurses’ attitudes toward telenursing.

A subset of telehealth, telenursing denotes the delivery, management, and coordination of care and services using telecommunication technologies by nurses [2]. Telenursing requires new nursing skills and competencies on the professional, methodological, personal, and social levels. Care delivery through audio calls, for example, requires greater interviewing competency, as nurses lose the possibility of using visual assessment for decision-making [3]. Hence, it is worthwhile that nurses learn and practice the fundamentals of remote communication with patients. Telenursing began with the advent of the telephone, as nurses handled healthcare questions over the phone. Over the last decade, the telenursing services have expanded beyond phone triage to videoconferencing. However, its implementation has raised concerns regarding costs, lack of data on safety, efficacy, and cost-effectiveness. Moreover, the provision of telenursing treatments without training has been shown to cause discomfort to both the nurse and the patient [4,5]. Thus, there is an evident need to prepare nurses for the new global challenges and demands of digital technology-based virtual healthcare provision [1,6].

The new training approach in nursing studies places the student at the center and the curriculum mainly focuses on problem-solving, simulations, and distant learning [7–9].

But training, too, has been shifting online. Mobile device-based education has been shown to vastly improve students’ learning motivation, self-confidence, communication competency, and perceived usefulness [10]. However, despite digital technology’s great advantages, barriers to using it for learning among nursing students still exist. Students and teachers alike still do not take full advantage of these platforms’ potential and have difficulty and poor confidence in using them. Therefore, digital literacy-targeted educational interventions need to be an integral part of foundational nursing studies, as it will improve nursing students’ baseline digital literacy before commencing clinical placement [8,11].

Nurses’ perceptions and acceptance of technology based on behavioral models are important as their view is associated with performance and may directly influence clinical outcomes and patient satisfaction [12,13]. The Theory of Planned Behavior (TPB) and the Technology Acceptance Model (TAM) are commonly used to assess the factors that contribute to acceptance of new technologies. According to TAM, the motivation for users’ acceptance is explained by a number of factors, including: (1) the degree to which a person believes that using a particular system would enhance his or her job performance (perceived usefulness, PU); (2) the degree to which a person believes that using a particular system would be free of effort (perceived ease of use, PEOU); and (3) an individual’s positive or negative feelings (evaluative affect) about using the technology (attitude toward use, AT) [14].

According to the TPB model, users’ technology acceptance depends on a number of predictors, including: (1) the perceived social pressures from important others encouraging them to perform the behavior (subjective norms, SN); (2) the person’s perceived ease or difficulty in performing the behavior (perceived behavioral control, PBC) [14]; and 3) the likelihood that a person will overcome potential obstacles (self-efficacy) [13].

Previous studies demonstrated that among healthcare workers, and specifically nurses, the intention to use a given technology was best predicted by the perceived ease of use, perceived social influence, and perceived usefulness for patient care. Also, training and support were found to be the most significant factors affecting the perceived ease of use and perceived usefulness [15–17]. Previous studies found that nurses’ attitudes towards telenursing correlated positively with their intention to participate in telenursing, with more positive attitudes expressed by nurses who were already using the technology or who perceived the technology to be simple and easy to use [18–23].

To the best of our knowledge, previous studies concerning perceptions towards adopting telenursing technologies did not compare the attitudes of nurses who underwent traditional frontal training before COVID-19 with those of nurses who trained online during or after COVID-19. Hence, the aim of this study was to explore behavioral-related factors based on both TAM and TPB for predicting the intention to use telenursing through videoconferencing and to compare those attitudes of nurses that undertook frontal (before COVID-19) vs. online (during and after COVID-19) post-basic nursing courses.

## 2. Methods

### 2.1. Study Design and Participants

We conducted a cross-sectional online survey among nurses working mainly at hospitals in Israel who undertook post-basic education training for nurses in the Dina academic nursing school at Rabin medical Center, Petah-Tikva, Israel, between January 2017 and December 2022. The survey was conducted between 6–28 December 2022.

Participants were recruited via convenience sampling, with a minimum overall target of 100 nurses per training method (traditional vs. online training). The inclusion criteria were nurses being trained in a post-basic education training in the Dina academic nursing school between January 2017 and December 2022, and 18 years of age and older. According to these criteria, the questionnaire was emailed to nurses that appeared on the appropriate lists.

At the beginning of the questionnaire form (Supplementary Questionnaire 1), the respondents were informed that their participation was voluntary and that they were permitted to terminate their participation at any time and were requested to confirm their informed consent to participate in the research. The interviewers followed a pre-defined closed-end protocol.

Participants who refused to give their consent to proceed with the questionnaire and those under the age of 18 years were excluded. The questionnaire was in Hebrew. The participants in the study received a full explanation of the structure and essence of the study and that it is anonymous.

### 2.2. Ethical Considerations

An exemption from Helsinki ethical approval was granted by the Helsinki Committee of Rabin Medical Center, Israel.

### 2.3. Questionnaire

The following sections describe the dependent and independent variables in the questionnaire and their operationalization in this study. The parameters comprising the study measurements used to build the conceptual model and hypotheses are described in Supplementary Figure 1.

The questionnaire consisted of the following sections: (A) TAM, TPB and accessibility to patient questions; (B) Telenursing experience and recommendation questions; and (C) socio-demographic and professional characteristics. The questionnaire was composed in total of 53 questions (the full questionnaire appears in Supplementary Questionnaire 1).

### 2.4. Measurement and Variables

The dependent variable was the intention to use telenursing, determined by 5 items measured on a 1–6 scale (1-strongly disagree, 6-strongly agree). User acceptance in this study was examined by the intention to use the technology, telenursing, via videoconference.

The independent variables were: 1). Socio-demographic and professional characteristics: (1) Age; (2) Gender; (3) Education; (4) Ethnicity; (5) Degree of religiosity and (6) Seniority. 2) TAM covariates: Perceived usefulness (PU), Perceived ease of use (PEOU) and Attitude (AT), measured by 6 items for each covariate. 3) TPB covariates: Subjective norms (SN), measured by 2 items, Perceived Behavioral Control (PBC), measured by 3 items, and Self-efficacy (SE), measured by 2 items. 4) Accessibility to patients, measured by three items. Accessibility to patients refers to the degree to which the nurse believes the use of the telenursing service will increase contact with underserved patients who live in regions that are geographically distant from medical facilities or who have difficulty getting to a medical facility.

All TAM, TPB and accessibility to patient items were measured on a 1–6 scale (1-strongly disagree, 6-strongly agree). Negative items were reverse scored, so that higher scores indicated higher levels of the item. Scores for each item were averaged to obtain each of the TAM and TPB independent categories.

To comprehend the perspective of nurses on the potential expansion of telenursing, three open-ended questions were formulated to explore their attitudes towards new areas it can be utilized in. The nurses who reported receiving training on communicating with remote patients were queried about their opinions regarding the beneficial aspects of the training and areas they believed could be enhanced.

### Reliability of the questionnaire

A Cronbach’s α internal reliability method revealed that the internal consistency of TAM was Cronbach’s α=0.89 and that of TPB was Cronbach’s α=0.83. The internal consistency of the integrated model was Cronbach’s α=0.87. (Table 1)

**Table 1.** Items, response scales and internal consistency for assessing measures of 2 models: TAM, TPB.

| Model | Measures | Items | Mean | Std | a |
| --- | --- | --- | --- | --- | --- |
| **TAM** | Attitude towards use (AT) | Using telenursing technology in patient care and management is a good idea. | 4.16 | 1.02 | .80 |
|  |  | Using telenursing technology in patient care and management is pleasant. |  |  |  |
|  |  | Using telenursing technology is beneficial to my patient care and management. |  |  |  |
|  |  | Using telenursing technology facilitates better patient care decision making. |  |  |  |
|  |  | *Using telenursing technology represents a violation of patient privacy. |  |  |  |
|  |  | *Using telenursing technology causes nurses to give less time to quality patient care. |  |  |  |
|  | Perceived usefulness (PU) | Using telenursing can enable me to complete patient care more quickly. | 3.96 | .96 | .91 |
|  |  | Using telenursing can improve my patient care and management. |  |  |  |
|  |  | Using telenursing can increase my productivity in patient care. |  |  |  |
|  |  | Using telenursing can enhance my service effectiveness. |  |  |  |
|  |  | Using telenursing can make my patient care and management easier. |  |  |  |
|  |  | I would find telenursing technology will be useful for my patient care and management. |  |  |  |
|  | Perceived ease of use (PEOU) | Learning to operate telenursing technology would be easy for me. | 4.49 | .89 | .91 |
|  |  | I would find it easy to get telenursing technology to do what I need to do in my patient care and management. |  |  |  |
|  |  | My interactions with telenursing technology would be clear and understandable. |  |  |  |
|  |  | I find telenursing technology flexible to interact with. |  |  |  |
|  |  | It is easy for me to become skillful in using telenursing technology. |  |  |  |
|  |  | I would find telenursing technology easy to use. |  |  |  |
|  | Intention to use (ITU) | I intend to use telenursing technology in my patient care and management when it becomes available in my department or hospital. | 4.03 | 1.15 | .95 |
|  |  | I intend to use telenursing technology to provide health care and management routinely. |  |  |  |
|  |  | Whenever possible, I intend to use telenursing technology in my patient care and management. |  |  |  |
|  |  | To the extent possible, I would use telenursing to do different things, clinical or nonclinical. |  |  |  |
|  |  | To the extent possible, I would use telenursing in my patient care and management frequently. |  |  |  |
| **TPB** | Subjective norms (SN) | Colleagues at my health care services work think that I should use telenursing technology. | 3.65 | 1.07 | .80 |
|  |  | My supervisors who are assessing my patient care and management think that I should use telenursing technology. |  |  |  |
|  | Perceived behavioral control (PBC) | I would have the ability to use telenursing technology in my patient care and management. | 4.07 | 1.02 | .86 |
|  |  | I would have the knowledge to make use of telenursing technology in my patient care and management. |  |  |  |
|  |  | I would have the resources to make use of telenursing technology in my patient care and management. |  |  |  |
|  | Self-efficacy (SE) | I could complete a job or a task using telenursing if I could call someone for help if I get stuck. | 4.43 | 1.07 | .83 |
|  |  | I could complete tasks in my work using the telenursing system if I receive the relevant training |  |  |  |
| **Patient centered care** | Accessibility to patient | Telenursing technology will allow me to treat patients who live at further distances. | 4.82 | .99 | .93 |
|  |  | Telenursing technology will allow me to be in contact with patients who seldom come to the clinic. |  |  |  |
|  |  | Telenursing technology will allow me to be in contact with patients who cannot be easily delivered to the clinic. |  |  |  |
| **No model** | - | I am willing to invest time in participating in training on the telenursing system at my place of work |  |  |  |

a Cronbach indicates the internal consistency: **TAM** a= 0.89 **TPB** a= 0.83
Items Response scale: 1-6 agreement
\* Negative items were reverse scored.

### 6.6 Statistical analysis

Data processing and analysis were done using SPSS for Windows (Version 28) software and the Process add-on for SPSS (Version 3.5).

We conducted a series of chi-squared tests to analyze the differences in categorical variables between nurses who learned the post-basic course in its traditional frontal format before 2020 and those who learned it in its remote format after 2020.

Additionally, we conducted t-tests to examine the differences in numeric variables between the two groups of nurses.

Interactions between dependent and independent discrete variables were examined using reliability tests, according to specific characteristics of the variables, using the standard reliability tests (t-test and chi-square test, according to the specific characteristics of the variables).

We analyzed variables to uncover correlations and factors that might be associated between the dependent variable (intention to use telenursing) and the independent variables (including socio-demographic factors and variables from the TAM, TPB and accessibility to patient).

Finally, we performed a multivariate analysis to investigate determinants of intention to use telenursing by using a multivariate ordinary least squares (OLS) regression analysis, which enabled us to examine the distinct effect of each independent variable on the dependent variables.

## 3. Results

### 3.1. Participant Characteristics

Overall, 195 respondents completed the online survey. Of the respondents, 30.3% (n=59) were male and 69.7% (n=136) were female. The average age of the participants was 37.3 (SD=7.5), and the average seniority was 8.9 years (SD=6.3). 70.8% (n=138) had a BA academic degree, and 29.2% (n=57) had a MA or higher academic degree. 37.4% (n=73) of the nurses in the study undertook post-basic education in a traditional, frontal format (before 2020; herein “frontal training group”) and 62.6% (n=122) in a remote format (after 2020; herein “remote training group”).

### 3.2. Univariate Analysis

Univariate tests were utilized to examine the variations in the demographics of the two groups. The seniority of those in the frontal training group (M=10.97, SD=4.85) was greater than those in the remote training group (M=7.73, SD=4.85) (t(193)=3.59, p<0.001, Cohen’s d=0.53). The age of nurses was found to have a close to significant difference in means. Specifically, the age of the nurses in the frontal training group (M=38.56, SD=6.48) was marginally greater than that of the nurses in the virtual training group (M=36.67, SD=8.04) (t(193)=1.52, p=0.07, Cohen’s d=0.23). Lastly, a nearly significant difference was found in education (χ2(1)=3.39,p=.07), where those in the frontal training group were more likely to have an MA or higher degree (37%) than a BA (24.6%), while those in the virtual training group were more likely to have a BA degree (75.4%) than an MA or higher degree (63%). See Table 2 for the full results.

**Table 2.** Socio-demographic characteristics of the research sample (n = 195).

| | All subjects<br>(n= 195) | | | | Before 2020<br>(n= 73) 0.374 | | | | After 2020<br>(n= 122) 0.626 | | | | t / $\chi^2$ | p-value | Effect size |
| --- | --- | --- | --- | --- | --- | --- | --- | --- | --- | --- | --- | --- | --- | --- | --- |
|  | M | SD | N | % | M | SD | N | % | M | SD | N | % |  |  |  |
| Age | 37.3 | 7.52 |  |  | 38.56 | 6.48 |  |  | 36.67 | 8.04 |  |  | 1.52 | 0.07 | 0.23 |
| Seniority | 8.94 | 6.3 |  |  | 10.97 | 4.85 |  |  | 7.73 | 6.76 |  |  | 3.59 | <.001 |  |
| Gender |  |  |  |  |  |  |  |  |  |  |  |  | 2.5 | 0.11 |  |
| * Male |  |  | 59 | 30.3% |  |  | 27 | 37.0% |  |  | 32 | 26.2% |  |  |  |
| * Female |  |  | 136 | 69.7% |  |  | 46 | 63.0% |  |  | 90 | 73.8% |  |  |  |
| Education |  |  |  |  |  |  |  |  |  |  |  |  | 3.39 | 0.07 |  |
| * BA |  |  | 138 | 70.8% |  |  | 46 | 63.0% |  |  | 92 | 75.4% |  |  |  |
| * MA or higher |  |  | 57 | 29.2% |  |  | 27 | 37.0% |  |  | 30 | 24.6% |  |  |  |
| Religion |  |  |  |  |  |  |  |  |  |  |  |  | 0.66 | 0.72 |  |
| * Jewish |  |  | 116 | 59.5% |  |  | 41 | 56.2% |  |  | 75 | 61.5% |  |  |  |
| * Muslim |  |  | 56 | 28.7% |  |  | 22 | 30.1% |  |  | 34 | 27.9% |  |  |  |
| * Other |  |  | 23 | 11.8% |  |  | 10 | 13.7% |  |  | 13 | 10.7% |  |  |  |
| Religiosity |  |  |  |  |  |  |  |  |  |  |  |  | 0.06 | 0.97 |  |
| * Secular |  |  | 82 | 42.1% |  |  | 31 | 42.5% |  |  | 51 | 41.8% |  |  |  |
| * Traditional |  |  | 63 | 32.3% |  |  | 24 | 32.9% |  |  | 39 | 32.0% |  |  |  |
| * Religious |  |  | 50 | 25.6% |  |  | 18 | 24.7% |  |  | 32 | 26.2% |  |  |  |
|  |  |  |  |  |  |  |  |  |  |  |  |  |  |  | 0.53 |

To test the bivariate correlations of all the TAM variables, TPB variables, accessibility to patient and the two continuous demographic variables (age and seniority) with the intention to use telenursing, all pairwise Pearson correlation coefficients were calculated. The results indicate that all TAM variables (attitude towards use, perceived usefulness, and perceived ease of use), all TPB variables (subjective norms, perceived behavioral control, and self-efficacy), and accessibility to patient were positively and significantly correlated with intention to use telenursing. The results suggest that when variables related to TAM or TPB increase, so does the intention to use telenursing.

Additionally, the accessibility to patients was found to have a positive correlation with the intention to use telenursing. However, the study did not find any correlation between age or seniority and the intention to use telenursing.

Furthermore, all TAM variables, TPB variables and accessibility to patient were significantly and positively correlated with one another. Lastly, age and seniority were also positively correlated with each another. See Table 3 for full results.

**Table 3:** Pearson correlations.

|  | M | SD | 1. | 2. | 3. | 4. | 5. | 6. | 7. | 8. | 9. |
| --- | --- | --- | --- | --- | --- | --- | --- | --- | --- | --- | --- |
| 1. Intention to use telenursing | 4.03 | 1.15 |  |  |  |  |  |  |  |  |  |
| 2. Attitude towards use | 4.17 | 1.03 | .70** |  |  |  |  |  |  |  |  |
| 3. Perceived usefulness | 3.97 | 0.96 | .69** | .78** |  |  |  |  |  |  |  |
| 4. Perceived ease of use | 4.50 | 0.9 | .50** | .42** | .49** |  |  |  |  |  |  |
| 5. Subjective norms | 3.65 | 1.07 | .71** | .56** | .59** | .34** |  |  |  |  |  |
| 6. Perceived behavioral control | 4.08 | 1.03 | .68** | .49** | .52** | .57** | .68** |  |  |  |  |
| 7. Self-efficacy | 4.44 | 1.07 | .53** | .42** | .43** | .36** | .52** | .61** |  |  |  |
| 8. Accessibility to patient | 4.82 | 1 | .54** | .56** | .56** | .47** | .50** | .60** | .66** |  |  |
| 9. Age | 37.3 | 7.52 | -0.05 | 0.00 | -0.08 | -0.09 | 0.01 | -0.09 | -0.04 | 0.00 |  |
| 10. Seniority | 8.94 | 6.3 | 0.04 | 0.06 | 0.02 | -0.09 | .15* | 0.00 | -0.03 | -0.04 | .62** |

\*\* . Correlation is significant at the 0.01 level (2-tailed).
\* . Correlation is significant at the 0.05 level (2-tailed).

To test differences in all TAM variables (attitude towards use, perceived usefulness and perceived ease of use), all TPB variables (subjective norms, perceived behavioral control and self-efficacy) and accessibility to patient among nurses in the frontal training group compared to nurses in the virtual training group, univariate tests were calculated. No significant differences were found in any of the tests (see Supplementary Table 1).

### 3.3. Multivariate Analysis

A multivariate OLS regression was calculated with intention to use telenursing as the dependent variable and with course time, demographic variables, TAM variables, TPB variables, and accessibility to patient as independent variables.

As is accepted, continuous variables were centered prior to the creation of an interaction term [24]. The final model indicated that the independent variables explained 71.4% of the variance in the intention to use telenursing (F(17,177)=26.05, P<0.001, R2=0.714, Adjusted R2=0.687). An examination of the model coefficients indicated a positive effect of both perceived ease of use and of subjective norms on the intention to use telenursing (β=0.29, p=0.03; β=0.61, p=0.00).

Beyond these main effects, a significant interaction term between subjective norms and course time was found to be a significant predictor of the intention to use telenursing (β= −0.33, p=0.03). To explain the interaction term, simple slopes were calculated using the Process v.4.2 add-on to SPSS[24]. The simple slope analysis revealed that in the frontal training group there was a positive relation between subjective norms and intention to use (b=0.53, se=0.12, β=0.49, p<0.001, 95%CI [0.29, 0.78]). A positive relation between subjective norms and intention to use was also found in the virtual training group, although this effect was weaker in magnitude (b=0.21, se=0.08, β=0.19, p=0.02, 95%CI [0.04, 0.37]). See Table 4 for a full model of the coefficients.

**Table 4.** Multivariate analyses of intention to use telenursing.

|  | B | SE | Beta | P-Value | 95%CI (LLCI) | 95%CI (ULCI) |
| --- | --- | --- | --- | --- | --- | --- |
| (Constant) | 3.83 | 0.22 |  | **<.001** | 3.40 | 4.26 |
| Course time | 0.12 | 0.10 | 0.05 | 0.26 | -0.09 | 0.32 |
| **Sociodemographic** |  |  |  |  |  |  |
| Seniority | 0.00 | 0.01 | 0.01 | 0.90 | -0.01 | 0.02 |
| Gender | 0.04 | 0.11 | 0.02 | 0.69 | -0.17 | 0.25 |
| **TAM** |  |  |  |  |  |  |
| Attitude towards use | 0.31 | 0.18 | 0.28 | 0.08 | -0.04 | 0.66 |
| Perceived usefulness | -0.13 | 0.21 | -0.11 | 0.52 | -0.54 | 0.28 |
| Perceived ease of use | 0.37 | 0.17 | 0.29 | **0.03** | 0.04 | 0.70 |
| **TPB** |  |  |  |  |  |  |
| Subjective norms | 0.65 | 0.17 | 0.61 | **0.00** | 0.31 | 0.99 |
| Perceived behavioral control | 0.15 | 0.17 | 0.13 | 0.39 | -0.19 | 0.49 |
| Self-efficacy | 0.01 | 0.13 | 0.01 | 0.94 | -0.25 | 0.27 |
| **Accessibility to patient** | -0.12 | 0.16 | -0.10 | 0.46 | -0.44 | 0.20 |
| Attitude towards use * Course | 0.02 | 0.16 | 0.02 | 0.90 | -0.30 | 0.34 |
| Perceived usefulness * Course | 0.31 | 0.18 | 0.28 | 0.09 | -0.05 | 0.67 |
| Perceived ease of use * Course | -0.23 | 0.15 | -0.20 | 0.11 | -0.52 | 0.05 |
| Subjective norms * Course | -0.33 | 0.15 | -0.33 | **0.03** | -0.62 | -0.03 |
| Perceived behavioral control * Course | 0.08 | 0.16 | 0.07 | 0.62 | -0.24 | 0.40 |
| self-efficacy * Course | 0.04 | 0.13 | 0.04 | 0.77 | -0.22 | 0.30 |
| Accessibility to patient * Course | 0.04 | 0.15 | 0.03 | 0.80 | -0.25 | 0.33 |

### Exploring the Pros and Cons of Telenursing Education for Nurses: Strategies for Enhancing Learning Outcomes

The nurses who had been trained in telenursing responded positively to questions regarding their opinions on the training program (e.g., “On the basis of your training experience, what were the positive aspects of training?). They further found the simulations and practice on a simulated patient to be helpful, and also the feedback their received from their instructor on their performance. In addition, they appreciated learning about potential clinical and technical issues that may arise and possible solutions to address them.

However, when asked about what they think should be improved in the training, they felt that the duration of the training program was insufficient and expressed a desire for more training time, increased patient simulation exercises, and access to written materials for future reference. Additionally, they suggested having an instructor provide personal support during the initial post-training period.

Table 5 presents the results of a thematic analysis conducted on the open-ended question, “For which purposes do you think telenursing technology could also serve, beyond triage?”. The responses led to the identification of three primary themes: (1) guidance and education for patients and families; (2) improving patient care assessments and patient follow-ups, and (3) counseling.

**Table 5:** The Potential Applications of Telenursing Technology Beyond Triage in Healthcare: main themes.

| Theme | Representative Quotations |
| --- | --- |
| **Guidance and education for patients and families** | “According to my perspective, telenursing has the potential to provide instruction in various areas, including, but not limited to, elective surgery or other procedures...”<br>“I think nurses can use this platform to guide patients on self-care in many fields, for example, instructions regarding taking care of a wound or a stoma or a gastric tube.”<br>“Video conferencing with a nurse can be beneficial and very useful in performing instructions in different topics, for example, instructions regarding medications, instructions regarding equipment, instructions regarding an insulin injection.”<br>“The use of telenursing can help the nurse demonstrate to the patient and his family how to perform various operations, such as connecting antibiotics to a PICC line or changing a urine bag in a patient with an indwelling catheter, or measuring blood glucose levels with a glucometer and even how to perform an injection.” |
| **Improving patient care assessments and patient follow-ups** | “I think telenursing has the potential to assist in the evaluation of patients who are homebound, as well as in conducting preliminary assessments prior to emergency room visits.”<br>“More ideas in which telenursing can help improving patients experience are assessment after surgery or after discharge from hospital, assessment and follow up of chronic wounds, proactive conversations to identify existing or potential problems, pregnancy monitoring and follow-up, and infant development follow-up.” |
|  | “Some follow-ups can be done by a nurse practitioner, which can help shorten queues and increase patient satisfaction.” |
| **Counseling** | “Telenursing is suitable for providing independent services such as sleep counseling, breastfeeding counseling, and nurse practitioner-led counseling on topics like insulin dosage”.<br>“It is possible to use telenursing through video calls for consultation with a nurse in certain situations as a substitute for a telephone call, so that the nurse and the patient can see each other and the nurse can see the patient and be impressed by his appearance, his breathing, the color of his skin, etc.” |

## Discussion

The use of distance learning tools and telehealth technologies is crucial in the field of nursing, especially in light of the recent COVID-19 pandemic. Given the significant role nurses play in patient communication through telehealth, gaining insight into their attitudes towards telenursing is crucial. To the best of our knowledge, this is the first study that compares to compare the intention to use telenursing through videoconferencing of nurses that were trained in a frontal format (before COVID-19) with nurses that were trained in an online format (during and after COVID-19). Additionally, the study identified factors that could predict nurses’ intention to use telenursing.

The present study found a high overall intention to use telenursing among participants. The following factors were found (using TPB and TAM) to significantly and positively correlated with this intention: subjective norms, perceived behavioral control, self-efficacy, attitude towards telenursing, its perceived usefulness, and perceived ease of use.

According to previous studies, perceived usefulness is one of the most important factors that predict physicians’ and nurses’ intentions to use telemedicine [15,22,23] and among the most frequent variables found to explain behavioral intention for using new technologies [16,17]. Our finding that perceived ease of use and subjective norms positively contribute to the intention to participate in telenursing via videoconference is consistent with a previous study that reported that ease of use was the only significant predictor of the behavioral intention to use technology among intensive care nurses [25]. Perceived ease of use has also been found to strongly influence the implementation process and to be greatly affected by training and management support. [15] [26]

Our finding that subjective norms, which reflects the influence of both peers and supervisors, contributes to nurses’ intention to practice telenursing is similar to that of Chang et al. [18], who suggested that promoting the development of telehealth services within an organization could encourage nurse participation in telenursing by organizing teams to work on technology solutions together. Barzeker et al. [15], too, identified management support, which includes staff training, technical support, and guidelines for technology systems, as crucial for nurses’ adoption of new technologies. Earlier research has shown that the perceived influence of co-workers [16,17,27] and the receipt of support from colleagues and the organization [26] both play a crucial role in determining nurses’ intention to use a new technology.

No significant differences were found between the two modes of training, despite the interaction between subjective norms and course time having a significant impact on nurses’ intention to use the technology. These findings suggest that the presence of supportive colleagues and supervisors who endorse the use of a technology is important for promoting its adoption among nurses, regardless of whether they were trained before or after the pandemic, or via traditional or online methods.

Based on the open-ended questions posed to nurses in the study, it can be inferred that even after undergoing telenursing training, the nurses still lacked confidence in utilizing the technology. This was largely due to their perception that they had not been provided with sufficient opportunities to practice.

We further identified, based on the nurses’ suggestions, three directions to which telenursing could be expanded: Guidance for patients and families, improving patient care assessments and follow-up, and counselling. The participants noted numerous areas in which telenursing can provide instruction, from elective surgery and other procedures, to medication administration and breastfeeding counseling.

Telenursing can provide greater accessibility to patients and families who may face geographical, financial, or mobility barriers in accessing in-person healthcare services. Indeed, a systematic review of providers’ attitudes toward tele mental health via videoconferencing concluded that its relative advantages, such as its ability to increase access to care, may outweigh its disadvantages [28]. Previous studies, however, merely examined this variable in the context of TPB and TAM models. Our finding that accessibility to patient was positively and significantly correlated with nurses’ intent to use telenursing via videoconference proves that nurses put the patient at the center and think about their best interests when considering the use of a new technology.

Telenursing requires nurses to possess not only a high level of clinical expertise but also the technological skills needed to operate the system. However, this can pose a challenge as nursing training programs often do not provide sufficient preparation in this area. Despite the significant benefits of telenursing and the positive attitudes towards it, there remains a gap between the level of training required for nurses to effectively use this technology and the level of preparation that currently exists. Furthermore, barriers still exist for nursing students seeking to become proficient in the digital technology required for telenursing, and only a few nursing education programs have integrated telehealth content into their curriculum [4].

The direct outcome is that the utilization of these tools is still very low, with nursing students often lacking the necessary knowledge and skills to use them effectively. For example, there is little focus on teaching students how to communicate remotely with patients. To address this issue, targeted digital literacy education interventions should be incorporated into foundational nursing studies. This can help improve nursing students’ baseline digital literacy and equip them with the skills needed to use telehealth technologies effectively in their practice. This education should include training on how to communicate with patients remotely, how to use telehealth equipment, and how to maintain patient privacy and confidentiality in a digital environment. Therefore, nursing educators and institutions should take an active role in incorporating digital literacy education and telehealth training into their curriculum [29] [8,11].

### Study limitations

It is important to acknowledge some limitations of this study. First, although the questionnaire was anonymous, there may still be a social desirability bias. Second, the study used a convenience sample, which may introduce bias and may not be representative of the entire population of nurses. The potential biases in this study include selection bias, researcher bias, and sampling bias. Finally, the study was conducted after the COVID-19 pandemic, during which time people had become accustomed to using various technologies and performing actions remotely. This could also affect the responses of nurses who were trained traditionally before the pandemic. Despite these limitations, our study provides valuable insights into the factors influencing nurses’ intention to practice telenursing. By examining nurses’ perceptions of telenursing, we were able to explain their willingness to use the technology, which is a precursor to their actual behavior.

## Conclusions

This study highlights the importance of considering nurses’ perceptions when introducing new technologies in healthcare, particularly after the changes brought about by the COVID-19 pandemic. The digital transformation of the healthcare sector requires not only technological changes but also adaptive changes in human attitudes and skills, legal frameworks, and work organization. We found that perceived ease of use and subjective norms were leading predictors of nurses’ intentions to use telenursing technology via videoconference, regardless of whether they learned about it through face-to-face or online training. Therefore, managers introducing telenursing should consider these factors and provide sufficient practical training to the nursing staff to ensure successful integration. Our results suggest that promoting telenursing technology can be done through educational courses and seminars, webinars, and support from direct managers and colleagues. This can help facilitate the adoption of the technology among nurses.

## Ethical Approval

The study was approved by the Ethics Committee for Non-clinical Studies at Rabin Medical Center, Israel.

## Data Availability

All data produced in the present study are available upon reasonable request to the authors

---

**Supplementary Figure 1:**
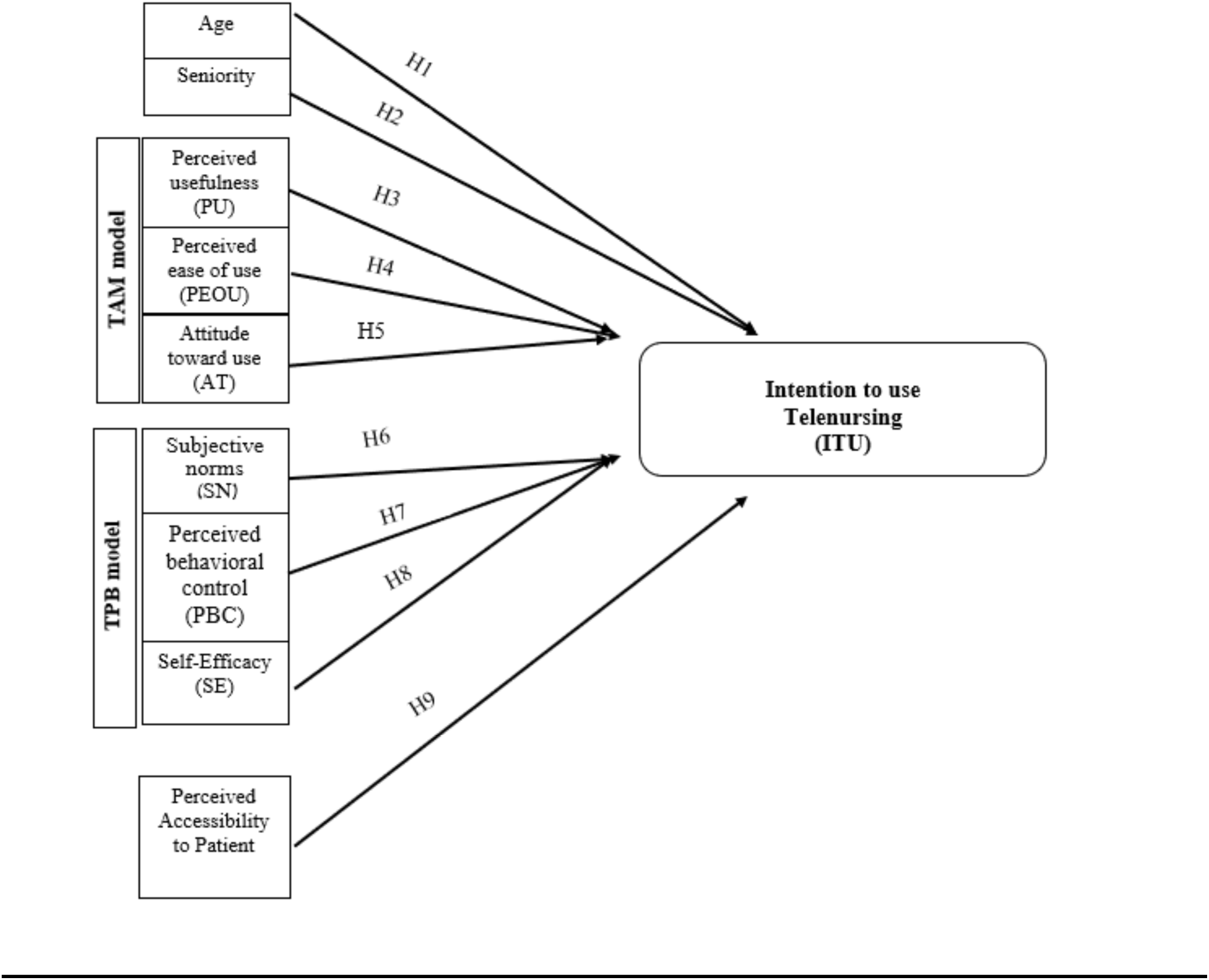
Research model and Hypotheses.

## Research Hypothesis

| **hypothesis** | **description** |
| --- | --- |
| H1 | Age negatively influences intentions to use telenursing. |
| H2 | Seniority negatively influences intentions to use telenursing. |
| H3 | Perceived usefulness positively influences intentions to use telenursing. |
| H4 | Perceived ease of use positively influences intentions to use telenursing. |
| H5 | Attitude positively influences intentions to use telenursing. |
| H6 | Subjective norms positively influence intentions to use telenursing. |
| H7 | Perceived behavioural control positively influences intentions to use telenursing. |
| H8 | Self-efficacy positively influences intentions to use telenursing. |
| H9 | Perceived accessibility to patient positively influences intentions to use telenursing. |
| H10 | All TAM variables will have interaction between course time on nurses' intention to use the technology. |
| H11 | All TPB variables will have interaction between course time on nurses' intention to use the technology. |

## Supplementary Questionnaire 1

### Questionnaire – Nurses’ Attitudes Towards the Use of “Telenursing”

Dear nurse,

The objective of the following questionnaire is to examine nurses’ attitudes to the use of “telenursing” (nurse-provided telemedicine services) through an electronic system for communicating with patients via remote video calls.

The questionnaire was conducted as part of a master’s thesis in health systems management at Bar-Ilan University. Please take a few minutes to complete this short, one-time questionnaire.

By filling out the questionnaire, you express your informed consent to participate in this study and declare that you are 18 years of age or older. You have the right to leave at any time without providing an explanation.

The questionnaire is anonymous and its use is intended for research purposes only. Thank you for participating in the study,

Svetlana Kats

1. 2. [A population-specific question]: How did you learn/are you learning the last post basic course at the ‘Dina’ academic nursing school?

1. Online only
2. Frontal instruction
3. Combined online and frontal instruction
4. did not take a basic course
2. What is your age?

Note: For respondents 18 years of age or younger, a form will be shown stating that the respondent is under 18 when filling the questionnaire.

Telenursing – definition:

Telenursing (a subdivision of telemedicine) refers to nurses’ provision of a medical service, provision of health information, prevention, and monitoring through an electronic platform/system for remote communication with patients (including phone and video calls and the use of remote monitoring equipment).

## Part A

The following statements refer to your attitudes towards the use of the “telenursing system” to communicate with patients via remote video calls. This system includes the use of a computer with access to the patient’s medical record and a video camera.

For each of the following items, please rate the degree to which you agree or disagree with the content of each statement, on a 1-6 scale:

1 – Strongly disagree
2 – Disagree
3 – Somewhat disagree
4 – Somewhat agree
5 – Agree
6 – Strongly agree

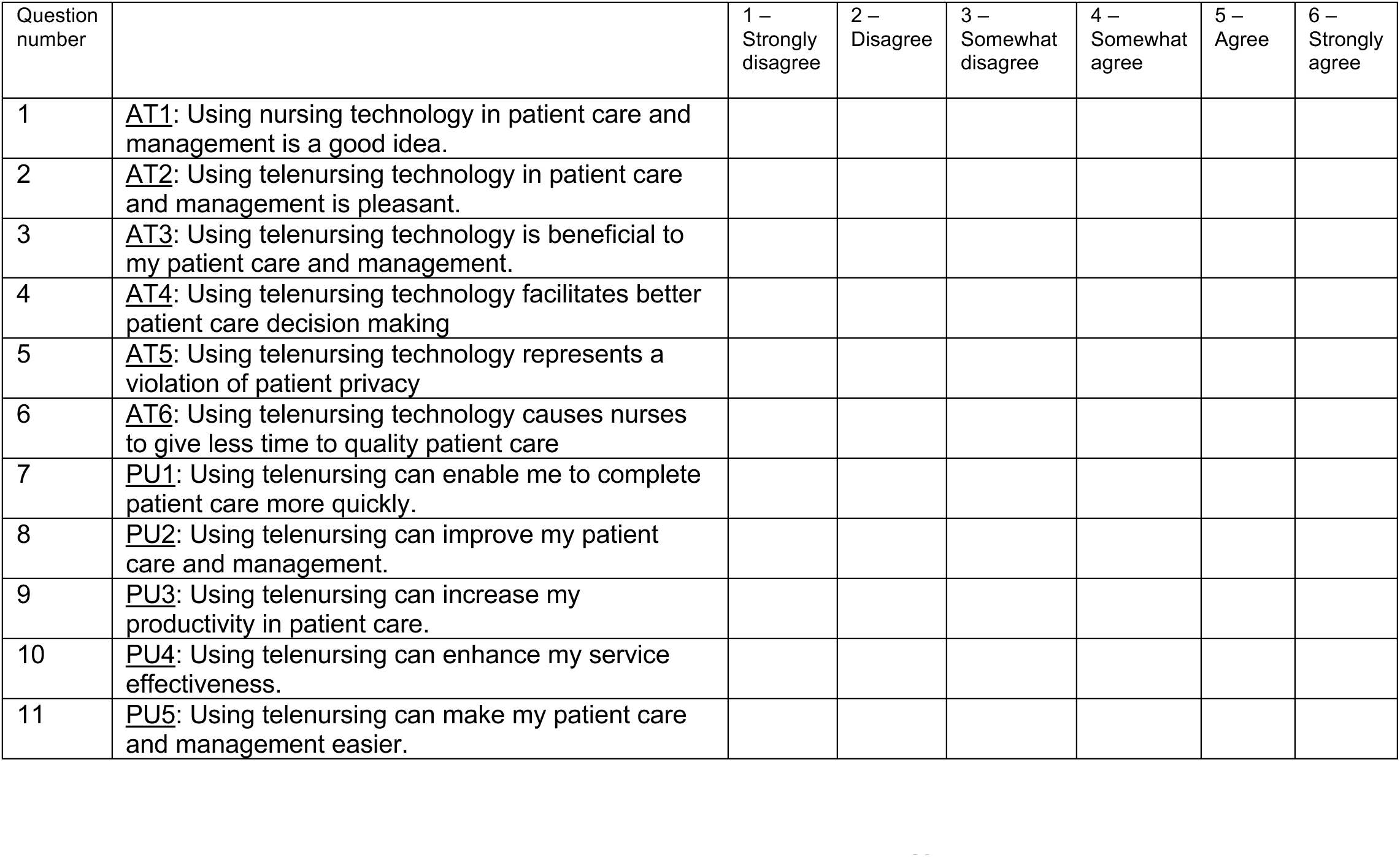
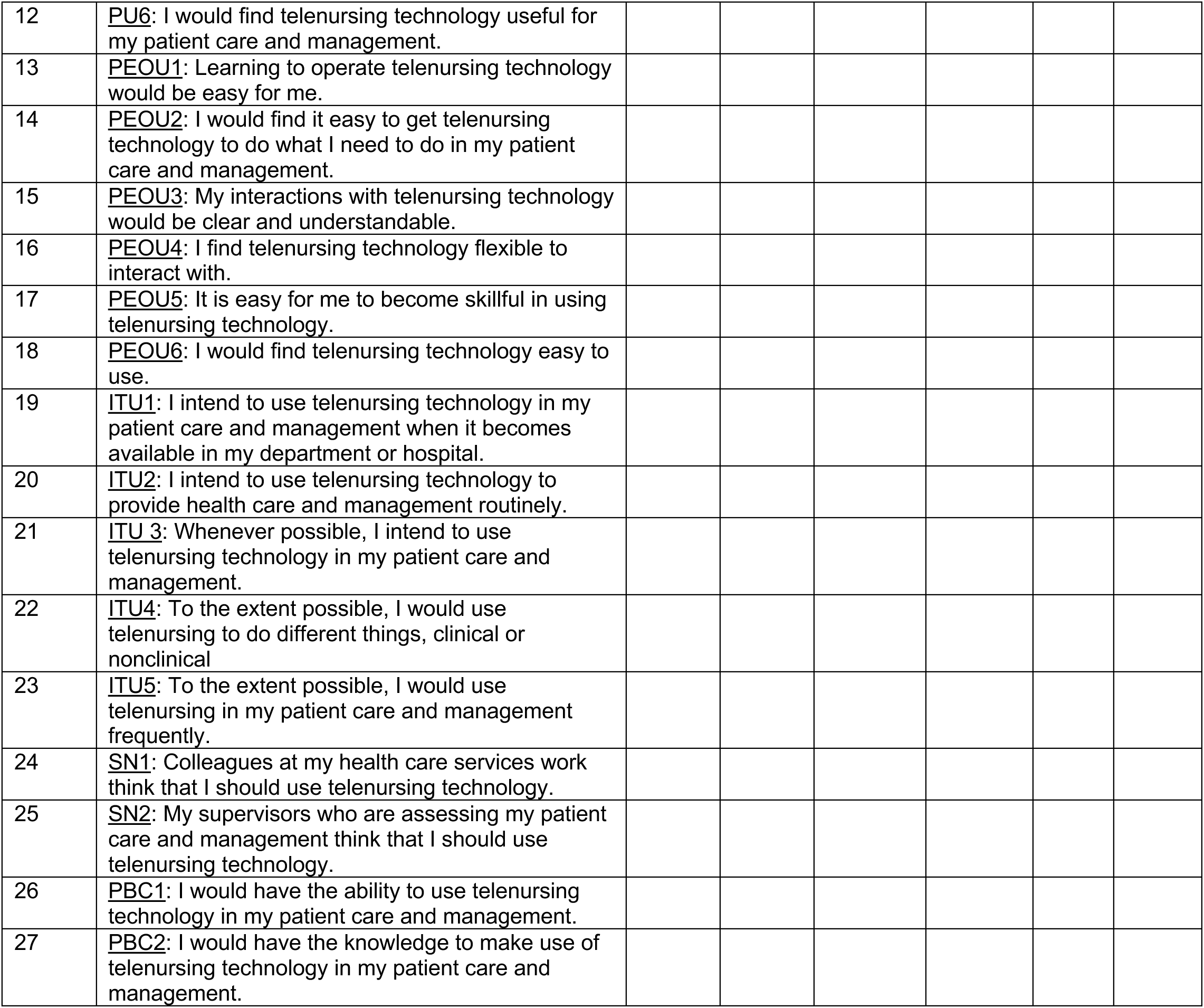
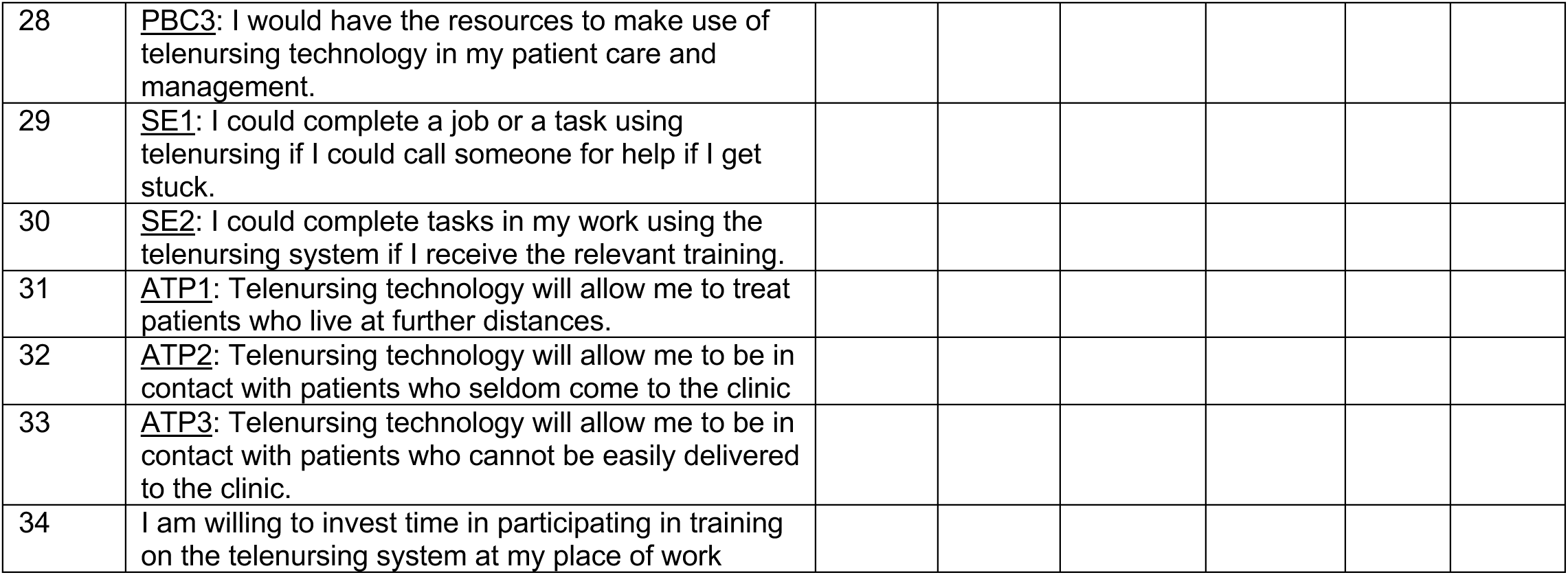

## Part B

1. Do you currently use a telernursing system to communicate with patients via video chats in your place of work?

1. Yes
2. No
2. Did you receive training on remote communication with patients via video chats, and if yes, in what form?

1. Yes, online training
2. Yes, frontal training
3. Yes, combined frontal and online
4. No
3. [If you answered Yes on question 2:] On the basis of your training experience,

1. What were the positive aspects of training? ______________________________
2. What do you think should be improved in the training? ______________________________
4. In what way would you recommend conducting training on the telenursing system?

1. Frontal instruction
2. An eLearning software
3. Online instruction in real time
4. Combined frontal and online instruction
5. Other – please specify: _________________
5. For which purposes do you think telenursing technology could also serve, beyond triage? ______________________________
6. When (in what year) did you last complete studies in a post basic education?

1. Before 2020, prior to the Covid pandemic
2. 2020 or later

## Part C

Please answer the following questions with regards to responder’s demographic background

1. What is your gender?

1. Male
2. Female
3. Other
2. What is your education? (The last school you attended)

1. Academic – bachelor’s degree
2. Academic – master’s degree or higher
3. What is your professional seniority as a nurse (number of years)? _______
4. What is your religion?

1. Jewish
2. Muslim
3. Christian
4. Druze
5. Other – please specify: _________________
5. How would you define your degree of religiosity?

1. Secular
2. Traditional
3. Religious
4. Devout/deeply religious
6. What is your primary place of work?

1. The community
2. A clinic/ institute in a hospital
3. Internal medicine department
4. Surgical department
5. Pediatric department
6. Intensive care
7. Operating room
8. Emergency medicine department (ER)
9. Other, please specify: _________________
7. What is your main work framework?

1. Public sector
2. Private sector
8. In which geographical area of Israel do you work?

1. Jerusalem district
2. The Northern District
3. Haifa district
4. Tel Aviv district
5. Central District
6. Southern District
7. Judea and Samaria area
9. What is your post basic education? Mark more than one answer as needed:

1. Clinical Instruction training
2. Operating room
3. adult intensive care
4. Emergency medicine
5. Pediatric intensive care
6. Neonatal intensive care
7. Nephrology
10. Are you a graduate of a clinical specialty program?

1. Yes, specify area: _________
2. No

## Supplementary Tables

**Supplementary Table 1:** Univariate analyses between TAM and TPB variables and the course.

|  | Before 2020<br>(n= 73) |  | After 2020<br>(n= 122) |  | t | p-value | Effect size |
| --- | --- | --- | --- | --- | --- | --- | --- |
|  | M | SD | M | SD |  |  | Cohen's d |
| **TAM** |  |  |  |  |  |  |  |
| Attitude towards use | 4.09 | 1.10 | 4.21 | 0.98 | 0.78 | 0.44 | 0.12 |
| Perceived usefulness | 3.96 | 1.00 | 3.97 | 0.94 | 0.03 | 0.98 | 0.00 |
| Perceived ease of use | 4.43 | 0.88 | 4.54 | 0.91 | 0.79 | 0.43 | 0.12 |
| **TPB** |  |  |  |  |  |  |  |
| Subjective norms | 3.71 | 1.12 | 3.62 | 1.05 | 0.52 | 0.61 | 0.08 |
| Perceived behavioral control | 3.95 | 1.13 | 4.16 | 0.95 | 1.39 | 0.17 | 0.21 |
| self-efficacy | 4.33 | 1.25 | 4.50 | 0.94 | 1.11 | 0.27 | 0.16 |
| Accessibility to patient | 4.68 | 1.09 | 4.91 | 0.93 | 1.56 | 0.12 | 0.23 |

## Bibliography

[1] Bell SA, Brysiewicz P. 2020 year of the nurse and midwife: Meeting new challenges. Int Emerg Nurs 2020;49:100848. https://doi.org/10.1016/j.ienj.2020.100848.

[2] Glinkowski W, Pawłowska K, Kozłowska L. Telehealth and Telenursing Perception and Knowledge Among University Students of Nursing in Poland. Telemed E-Health 2013;19:523–9. https://doi.org/10.1089/tmj.2012.0217.

[3] Fincher L, Ward C, Dawkins V, Magee V, Willson P. Using Telehealth to Educate Parkinson’s Disease Patients About Complicated Medication Regimens. J Gerontol Nurs 2009;35:16–24.

[4] Rutledge CM, Gustin T. Preparing Nurses for Roles in Telehealth: Now is the Time! Online J Issues Nurs 2021;26:1–13. https://doi.org/10.3912/OJIN.Vol26No01Man03.

[5] Chan WM, Hjelm NM. The role of telenursing in the provision of geriatric outreach services to residential homes in Hong Kong. J Telemed Telecare 2001;7:38–46. https://doi.org/10.1258/1357633011936129.

[6] Kumara WGC, Sudusinghe WC. Improving Nursing Education in Sri Lanka to take on New Challenges faced by Global Healthcare Systems. Univ Colombo Rev 2021;2:119. https://doi.org/10.4038/ucr.v2i1.42.

[7] Leong FF, He H-G, Premarani K, Lim SH. Nurses’ perceptions of nursing education transformation and its impact on care delivery in Singapore. Int Nurs Rev 2022;n/a. https://doi.org/10.1111/inr.12733.

[8] Lokmic-Tomkins Z, Choo D, Foley P, Dix S, Wong P, Brand G. Pre-registration nursing students’ perceptions of their baseline digital literacy and what it means for education: A prospective COHORT survey study. Nurse Educ Today 2022;111:105308. https://doi.org/10.1016/j.nedt.2022.105308.

[9] Purnell M, Royal B, Warton L. Supporting the development of information literacy skills and knowledge in undergraduate nursing students: An integrative review. Nurse Educ Today 2020;95:104585. https://doi.org/10.1016/j.nedt.2020.104585.

[10] Shanmugapriya K, Seethalakshmi A, Zayabalaradjane Z, Rani NRV. Mobile technology acceptance among undergraduate nursing students instructed by blended learning at selected educational institutions in South India. J Educ Health Promot 2023;12:45. https://doi.org/10.4103/jehp.jehp_488_22.

[11] Loureiro F, Sousa L, Antunes V. Use of Digital Educational Technologies among Nursing Students and Teachers: An Exploratory Study. J Pers Med 2021;11:1010. https://doi.org/10.3390/jpm11101010.

[12] Taherdoost H. Importance of Technology Acceptance Assessment for Successful Implementation and Development of New Technologies. Glob J Eng Sci 2019;1.

[13] Ajzen I. The theory of planned behavior: Frequently asked questions. Hum Behav Emerg Technol 2020;2:314–24. https://doi.org/10.1002/hbe2.195.

[14] Venkatesh V, Morris MG, Davis GB, Davis FD. User Acceptance of Information Technology: Toward a Unified View. MIS Q 2003;27:425–78. https://doi.org/10.2307/30036540.

[15] Barzekar H, Ebrahimzadeh F, Luo J, Karami M, Robati Z, Goodarzi P. Adoption of Hospital Information System Among Nurses: a Technology Acceptance Model Approach. Acta Inform Medica 2019;27:305–10. http://dx.doi.org/10.5455/aim.2019.27.305-310.

[16] Holden RJ, Brown RL, Scanlon MC, Karsh B-T. Modeling nurses’ acceptance of bar coded medication administration technology at a pediatric hospital. J Am Med Inform Assoc 2012;19:1050–8. https://doi.org/10.1136/amiajnl-2011-000754.

[17] Hung S-Y, Tsai JC-A, Chuang C-C. Investigating primary health care nurses’ intention to use information technology: An empirical study in Taiwan. Decis Support Syst 2014;57:331–42. https://doi.org/10.1016/j.dss.2013.09.016.

[18] Chang M-Y, Kuo F-L, Lin T-R, Li C-C, Lee T-Y. The Intention and Influence Factors of Nurses’ Participation in Telenursing. Informatics 2021;8:35. https://doi.org/10.3390/informatics8020035.

[19] Bashir A, Bastola DR. Perspectives of Nurses Toward Telehealth Efficacy and Quality of Health Care: Pilot Study. JMIR Med Inform 2018;6:e35. https://doi.org/10.2196/medinform.9080.

[20] Rho MJ, Choi I young, Lee J. Predictive factors of telemedicine service acceptance and behavioral intention of physicians. Int J Med Inf 2014;83:559–71. https://doi.org/10.1016/j.ijmedinf.2014.05.005.

[21] Napitupulu D, Yacub R, Halim Perdana Kusuma A. Factor Influencing of Telehealth Acceptance During COVID-19 Outbreak: Extending UTAUT Model. Int J Eng Intell Syst Electr Eng Commun 2021;14:2021. https://doi.org/10.22266/ijies2021.0630.23.

[22] Hu PJ, Chau PYK, Sheng ORL, Tam KY. Examining the Technology Acceptance Model Using Physician Acceptance of Telemedicine Technology. J Manag Inf Syst 1999;16:91–112. https://doi.org/10.1080/07421222.1999.11518247.

[23] Saigi-Rubió F, Jiménez-Zarco A, Torrent-Sellens J. DETERMINANTS OF THE INTENTION TO USE TELEMEDICINE: EVIDENCE FROM PRIMARY CARE PHYSICIANS. Int J Technol Assess Health Care 2016;32:29–36. https://doi.org/10.1017/S0266462316000015.

[24] Hayes AF. Introduction to mediation, moderation, and conditional process analysis: a regression-based approach. New York: The Guilford Press; 2013.

[25] Wilson R, Duhn L, Gonzalez P, Hall S, Chan YE, VanDenKerkhof EG. Wireless Communication in Clinical Environments with Unique Needs. J Healthc Qual 2014;36:24–32. https://doi.org/10.1111/jhq.12030.

[26] Konttila J, Siira H, Kyngäs H, Lahtinen M, Elo S, Kääriäinen M, et al. Healthcare professionals’ competence in digitalisation: A systematic review. J Clin Nurs 2019;28:745–61. https://doi.org/10.1111/jocn.14710.

[27] Zhang H, Cocosila M, Archer N. Factors of Adoption of Mobile Information Technology by Homecare Nurses: A Technology Acceptance Model 2 Approach. CIN Comput Inform Nurs 2010;28:49–56. https://doi.org/10.1097/NCN.0b013e3181c0474a.

[28] Connolly. A systematic review of providers’ attitudes toward telemental health via videoconferencing - Connolly - 2020 - Clinical Psychology: Science and Practice - Wiley Online Library 2020. https://onlinelibrary.wiley.com/doi/abs/10.1111/cpsp.12311 (accessed May 6, 2023).

[29] El-Said Abd Ellatif A, Mohamed Sobhy Elsayed D, Hamido Abosree T. Knowledge and Attitude of Faculty of Nursing Students regarding Telenursing. J Nurs Sci Benha Univ 2023;4:677–89. https://doi.org/10.21608/jnsbu.2023.278954.

